# Intranasal oxytocin improves interoceptive accuracy and heart-beat evoked potentials in a cardiac interoceptive task

**DOI:** 10.1101/2024.01.25.24301761

**Authors:** Menghan Zhou, Lanqing Cheng, Yibo Zhou, Siyu Zhu, Yuan Zhang, Keith M. Kendrick, Shuxia Yao

## Abstract

**Background:** Interoception represents perception of the internal bodily state which is closely associated with social/emotional processing and physical health in humans. Understanding the mechanism underlying interoceptive processing, particularly its modulation, is thus of great importance. Given overlap between oxytocinergic pathways and interoceptive signaling substrates in both peripheral visceral organs and the brain, intranasal oxytocin administration is a promising approach for modulating interoceptive processing.

**Methods:** In a double-blind, placebo-controlled, between-subject design, 80 healthy male participants were recruited to perform a cardiac interoceptive task during electroencephalograph (EEG) and electrocardiograph (ECG) recording to examine whether intranasal administration of the neuropeptide oxytocin can modulate interoceptive processing. We also collected data in a resting state to examine whether we could replicate previous findings.

**Results:** Results showed that in the interoceptive task oxytocin increased interoceptive accuracy at the behavioral level which was paralleled by larger heartbeat-evoked potential amplitudes on the neural level. Heartbeat-evoked potential amplitudes were found to be positively correlated with interoceptive accuracy across groups. However, there were no significant effects of oxytocin on EEG or ECG during resting-state.

**Conclusions:** These findings suggest that oxytocin may only have a facilitatory effect on interoceptive processing during task-based conditions. Our findings not only provide new insights into the modulation of interoceptive processing via targeting the oxytocinergic system but also provide proof of concept evidence for the therapeutic potential of intranasal oxytocin in mental disorders with dysfunctional interoception.

**Clinical Trials Registration:** Registry name: UESTC-neuSCAN-83 URL: https://register.clinicaltrials.gov/prs/app/action/ViewOrUnrelease?uid=U0002QSK%ts=14%sid=S000BB9A%cx=-xxsuzb

Registration number: NCT05245708

## Introduction

Interoception refers to perception of the internal state of one’s own body including cardiac, hunger, temperature, pain and respiratory signals (1,2). It plays an important role in social and high-level cognitive processing, including empathy, emotional memory, learning and decision-making (3–8). Interoceptive dysfunction has been associated with psychiatric disorders including anxiety, depression, addiction and alexithymia (9,10,2) as well as physical health problems, such as obesity and diabetes (11,12). Thus, understanding the mechanism underlying interoceptive processing, and particularly whether it can be modulated, is of great importance.

Oxytocin (OT) is a hypothalamic neuropeptide that has been shown to have numerous modulatory effects on social behaviors and emotional processing in both animals and humans (13,14), although direct evidence for its effects on interoceptive processing is limited. While OT receptors are distributed widely in visceral organs such as the heart and stomach, which are afferent hubs of interoceptive signals, the interoceptive neural pathway, including the insula and dorsal anterior cingulate cortex, also overlaps with brain networks underpinning OT’s regulatory effects on human behavior (15–17). It has therefore been proposed that OT can affect the interoceptive signal transmission of almost every modality and provide information about the internal environment related to homeostasis (18). In humans, findings of OT’s effects on interoceptive processing are divergent, depending on the paradigms used in different studies. Betka et al. (19) found that in alcohol users, while intranasal OT reduced interoceptive accuracy (IAc) in a heartbeat counting task (HCT) it increased IAc measured in a heartbeat discrimination task. Using a revised heartbeat detection task whereby participants made a button press every time they felt a heartbeat, OT was found to have no significant effects on IAc, but when participants were simultaneously presented with social stimuli, OT decreased IAc, possibly due to OT acting to switch attention away from interoceptive signals towards external social stimuli (20). These divergent findings could be due to that the way IAc being measured in these cardiac interoceptive paradigms is highly dependent on individuals’ ability to subjectively perceive their heartbeat, which may introduce confounding effects from time estimation and prior knowledge of heartbeat (21–23). Thus, a more objective index is preferable for examining OT’s effects on interoceptive processing and the heartbeat-evoked potential (HEP) could be a promising alternative.

The HEP is a scalp event-related potential that reflects cortical processing of cardiac interoceptive signals (24). It is usually locked in a time window of 200-600 ms after R waves at frontal, frontocentral and central electrodes (1,25–27) and has been proposed as a neurophysiological marker for interoception (1,28). Using the HCT, Montoya et al. (27) found that the HEP amplitude was significantly larger in good heartbeat perceivers relative to poor ones. Pollatos and Schandry (24) confirmed this finding using the same HCT and further found a positive correlation between the cardiac IAc scores and the HEP amplitude. However, until now there is only one study that has investigated OT’s effects on the HEP and reported that OT had no significant effects on resting-state HEP responses in both healthy individuals and those with borderline personality disorder (29). This still leaves open the possibility that intranasal OT can modulate the HEP when participants are performing a cardiac interoceptive task.

The present study therefore combined electrocardiograph (ECG) and electroencephalograph (EEG) recordings in the HCT to investigate whether OT could modulate the HEP as an objective neurophysiological marker for interoceptive processing. Based on previous findings of close associations between the HEP and empathy and emotional recognition (3,30), as well as findings of OT facilitating empathy and emotion recognition (31,32), we hypothesized that OT would increase amplitudes of the HEP compared with placebo (PLC). For behavioral responses, two indices were collected with IAc measuring individuals’ ability to perceive internal cardiac signals and interoceptive sensibility (IS) measuring their confidence for IAc (33). In the same vein as our hypotheses for the HEP, we predicted an enhancement effect of OT on interoceptive processing at the behavioral level (but see Betka et al., 2018). We also collected data in a resting state to examine whether we could replicate previous findings (29).

## Materials and Methods

### Participants and treatment

Eighty healthy male participants (mean age = 20.65 years, SD = 1.77) were recruited for the present randomized, double-blind, PLC-controlled, between-subject pharmacological study. The sample size was adequate to achieve a power of > 90% (effect size = 0.8, α = 0.05) for an independent *t*-test analysis based on a priori power analysis using the G*Power v.3.1 toolbox (34). Participants were all free of current or past psychiatric, neurological or other medical conditions. They were instructed to abstain from smoking, alcohol and caffeine for 24 hours and not to have any food or drinks except water for 2 hours prior to the experiment. Participants who reported great difficulty in feeling their heartbeat and perceiving it based on prior knowledge of their heartbeat were excluded (6 participants). Another 2 participants were excluded due to technical problems during data recording (1 participant) or as an outlier in the EEG data (1 participant). Consequently 37 participants in the OT group and 35 participants in the PLC group were included in final data analyses. To control for potential confounding effects from mood changes and personality traits, participants completed Chinese versions of validated questionnaires before treatment, including the Autism Spectrum Quotient (35), State-Trait Anxiety Inventory (36), Beck Depression Inventory (37,38), and Behavioral Inhibition System and Behavioral Activation System Scale (39). The Positive and Negative Affect Schedule (40) was completed 3 times: when participants arrived (pre-treatment), 45 minutes after treatment but before the task (post-treatment) and immediately after completing the task (post-task) to check for changes in mood during the whole experiment.

Participants were randomly assigned into two groups (OT vs. PLC) and self-administered either OT (OT-spray, Sichuan Defeng Pharmaceutical Co. Ltd, China) or PLC (identical ingredients with the OT-spray but without OT, i.e., sodium chloride and glycerin) nasal spray. 24 international units (IU) of OT or PLC were administered with 3 puffs to each nostril following a standardized protocol (41). 45 minutes after treatment, participants first performed an associative learning task (duration: 20 minutes; reported elsewhere) followed by a resting-state recording (4 minutes) and the HCT (around 3 minutes) while undergoing EEG and ECG measurements (see Fig. 1A). All participants provided written informed consent before the experiment. All procedures conformed with the latest version of the Declaration of Helsinki and were approved by the ethical committee of the University of Electronic Science and Technology of China.

**Fig. 1.**
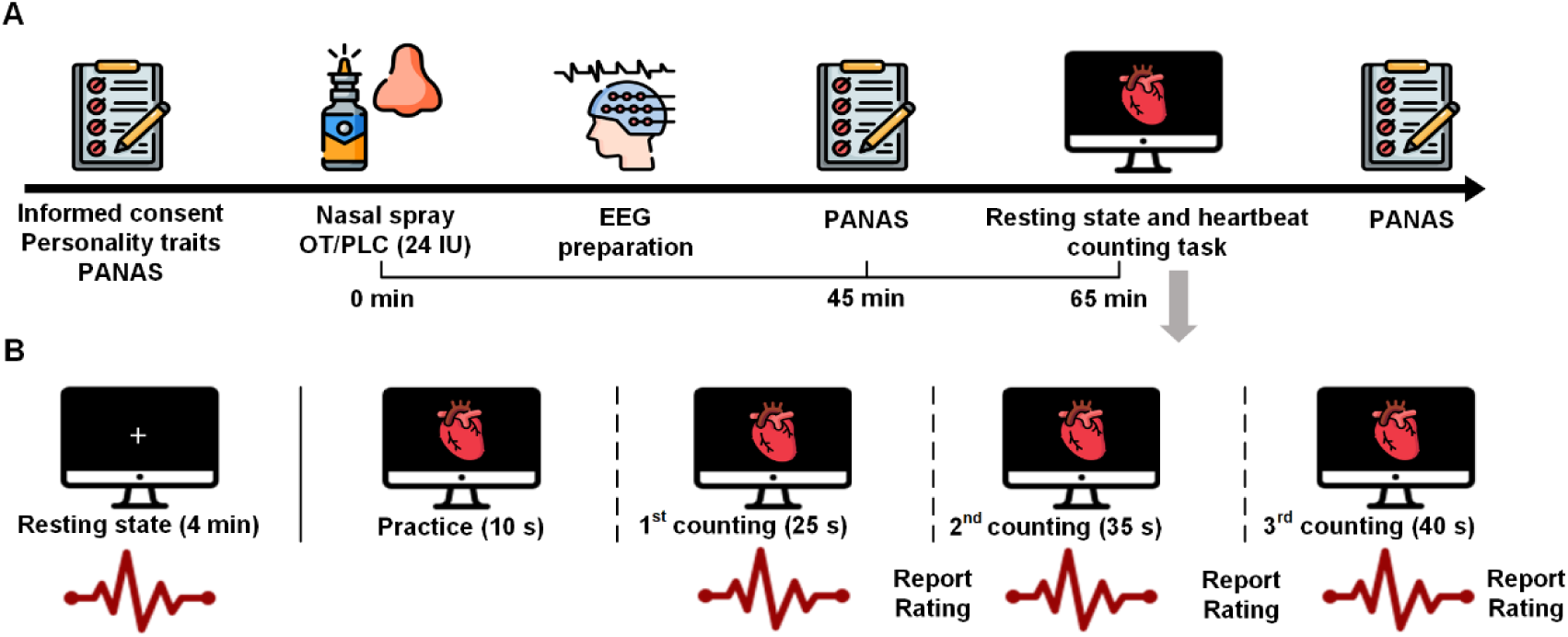
(A) Experimental protocol. (B) Timeline of the resting-state recording and the heartbeat counting task. License: Icons were obtained from flaticon.com under the free license with attribution.

### Experimental task

During the resting-state recording, participants were asked to sit comfortably and look at a white cross fixation against a black background on the computer monitor for 4 minutes (see also Al et al., 2021; Schmitz et al., 2020). The HCT was developed based on Schandry (43), in which participants were instructed to focus attention on their own heartbeats and count times of heartbeats silently (see Fig. 1B). Optimized instruction was used to minimize confounding effects from time estimation and prior knowledge of heartbeat (21,22). Participants were clearly informed that it was not allowed to use other methods such as by feeling their pulse as further aids for heartbeat detection. The HCT consisted of one practice session (10 s) and three formal sessions (25 s, 35 s and 40 s). Participants had no idea about the length of each counting session. The start and stop of each heartbeat counting session were signaled by the appearance and disappearance of a heart symbol. When a white fixation cross was presented after each counting session, participants were asked to verbally report the number of their counted heartbeats. Participants were then instructed to rate their level of confidence in their heartbeat perception (i.e., IS). IAc was calculated using the following formula:

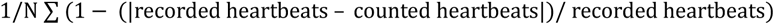

Where N = 3 corresponds to the number of counting sessions. Higher scores (maximum = 1) indicate higher accuracy in perceiving heartbeats. IS was calculated by the average of confidence ratings after the three sessions.

### ECG data acquisition and analyses

The ECG data was recorded using a BIOPAC MP150 system (BIOPAC Systems, Inc.; ECG100C module) at a sampling rate of 1000 Hz. Two electrodes were placed on participants’ lower left leg and the right wrist. The ECG data was processed using the AcqKnowledge software (Version 4.4, Biopac Systems Inc., CA, USA) in accordance with the manual. The raw data were band-pass filtered (0.5-35 Hz) to remove baseline drift and high-frequency noise. All data were manually inspected and data quality was high. Time points of R waves were detected using the find circle function and confirmed by visual inspection. R-R intervals were then extracted and imported into the Kubios software (http://kubios.uku.fi) for high-frequency heart rate variability (HRV) analyses.

### EEG data acquisition and analyses

The EEG was recorded at a sampling rate of 500 Hz using a 64-channel ActiCap system with a Quick Amp amplifier (Brain Products GmbH, Germany). Signals of all channels were online referenced to the Cz electrode with the ground on a medial prefrontal electrode (the international 10-20 system). Electrode impedances were kept below 5 kΩ. Offline EEG data was analyzed using the EEGLAB 14.1.1 toolbox (44). The raw EEG data was down-sampled to 250 Hz and filtered with a Hamming windowed sinc FIR filter separately for high- and low-pass filters (high-pass: 0.1 Hz, -6 dB cutoff: 0.05 Hz; low-pass: 40 Hz, -6 dB cutoff: 45 Hz). After re-referencing to the average reference, an independent component analysis (ICA) was conducted to reject components of eye movement, muscle and cardiac artifacts. To remove residual artifacts, epochs with voltage values exceeding ± 80 µV were further deleted. An average of 11.07% of trials were excluded from further analyses.

For the HEP, EEG data was extracted from 200 ms before and 800 ms after the peak of R waves. Based on previous studies (1,26), a time window of 480-600 ms was used to calculate the mean HEP amplitude at frontocentral and central electrodes (FC1, FCz, FC2, FC4, C1, Cz, C2, C4), two locations previously reported to show strong HEP responses (1,24,26,45).

### Statistical analyses

Independent *t*-tests were conducted to compare group differences on questionnaire scores, IAc and IS. For physiological and neural data, independent *t*-tests were also performed on the heart rate (HR), HRV and mean amplitude of the HEP in the resting-state and task conditions respectively. Furthermore, correlations between behavioral, physiological and neural measurements were analyzed using Pearson or Spearman correlations depending on distribution of the data. Correlation differences between treatments were tested using the Fisher z-transformation test.

## Results

### Demographics and questionnaires

Independent *t*-tests on personality traits revealed no significant group differences (see Table 1). There were also no significant group differences of both pre- and post-treatment measures of mood (see Table 2).

**Table 1.** Statistics of age and questionnaire scores of personality traits.

| Measurements | OT (M ± SD) | PLC (M ± SD) | *t*-values | *p*-values |
| --- | --- | --- | --- | --- |
| Age | 20.59±1.64 | 20.57±1.85 | 0.056 | 0.955 |
| Autism Spectrum Quotient (ASQ) | 23.41±5.32 | 21.31±5.17 | 1.691 | 0.095 |
| State-Trait Anxiety Inventory |  |  |  |  |
| -Trait Anxiety Inventory (TAI) | 42.00±7.37 | 42.51±8.70 | -0.271 | 0.787 |
| -State Anxiety Inventory (SAI) | 38.84±6.64 | 40.69±8.96 | -0.998 | 0.322 |
| Beck Depression Inventory (BDI-II) | 8.54±6.94 | 9.06±7.83 | -0.297 | 0.768 |
| Behavioral Inhibition System and Behavioral Activation System Scale |  |  |  |  |
| -Behavioral Inhibition System (BAS) | 22.95±5.07 | 24.77±4.42 | -1.625 | 0.109 |
| -Behavioral Activation System (BIS) | 14.95±3.34 | 15.43±2.68 | -0.674 | 0.503 |

Values are presented as mean ± SD. OT: oxytocin; PLC: placebo.

**Table 2.** Statistics of the Positive and Negative Affect Schedule scores.

|  | OT (M ± SD) | PLC (M ± SD) | *t*-values | *p*-values |
| --- | --- | --- | --- | --- |
| Positive and Negative Affect Schedule<sup>pre-treatment</sup> |  |  |  |  |
| -Positive | 25.86±6.37 | 25.94±6.43 | -0.052 | 0.959 |
| -Negative | 14.38±4.74 | 16.74±7.01 | -1.684 | 0.097 |
| Positive and Negative Affect Schedule<sup>post-treatment</sup> |  |  |  |  |
| -Positive | 23.57±7.30 | 23.31±7.37 | 0.147 | 0.884 |
| -Negative | 13.32±5.49 | 13.11±4.54 | 0.176 | 0.861 |
| Positive and Negative Affect Schedule<sup>post-task</sup> |  |  |  |  |
| -Positive | 22.08±6.78 | 23.97±7.05 | -1.160 | 0.250 |
| -Negative | 11.84±4.74 | 11.89±3.45 | -0.049 | 0.961 |

Values are presented as mean ± SD. OT: oxytocin; PLC: placebo.

### Resting-state condition

#### HR and HRV

Independent *t*-tests on HR and HRV revealed no significant group differences in the resting-state condition (*ps* ≥ 0.164).

#### HEP

To examine whether there was an effect of OT on the resting-state HEP, mean HEP amplitudes were extracted for each participant. An independent *t*-test showed no significant group difference of the mean HEP amplitudes (0.22 ± 0.44 vs 0.07 ± 0.44, *t*(70) = 1.395, *p* = 0.167).

### Task-based condition-HCT

#### HR and HRV

Independent *t*-tests revealed no significant group differences for HR and HRV in the task-based condition (*ps* ≥ 0.285).

#### IAc and IS

Independent *t*-tests were employed to analyze group differences of IAc and IS in the HCT and showed that IAc in the OT group was significantly higher than in the PLC group (0.77 ± 0.15 vs 0.68 ± 0.19, *t*(70) = 2.264, *p* = 0.027, Cohen’s d = 0.53; Fig. 2A). However, no significant group difference was found for IS (5.86 ± 1.48 vs 5.92 ± 1.57, *t*(70) = -0.165, *p* = 0.870; Fig. 2B).

**Fig. 2.**
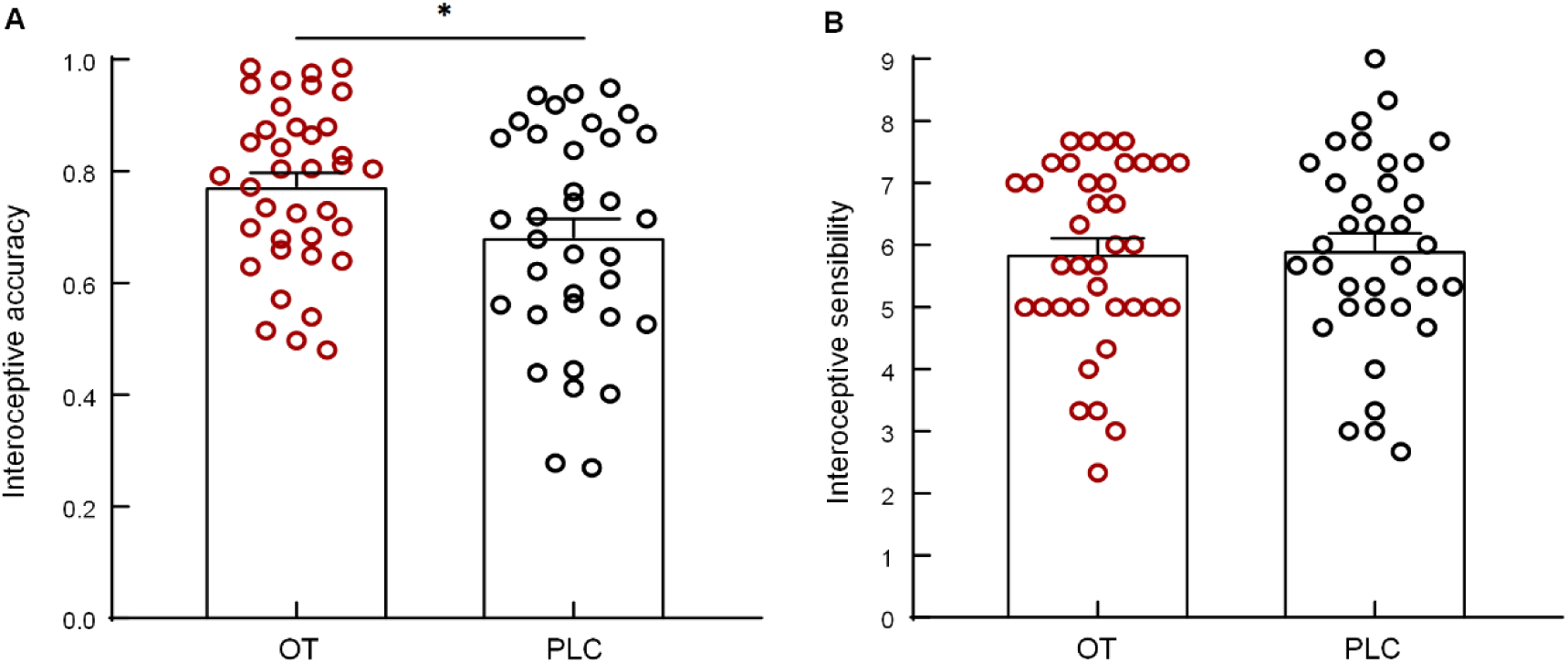
Interoceptive accuracy (A) and sensibility (B) in the OT and PLC groups measured in the heartbeat counting task.

#### HEP

To examine whether there was an effect of OT on the HEP in the HCT, mean HEP amplitudes were calculated for each participant. An independent *t*-test showed that the group difference in mean HEP amplitudes was significant with HEP amplitudes in the OT group being larger than in the PLC group (0.26 ± 0.41 vs 0.04 ± 0.30, *t*(70) = 2.650, *p* = 0.011, Cohen’s d = 0.61; Fig. 3).

**Fig. 3.**
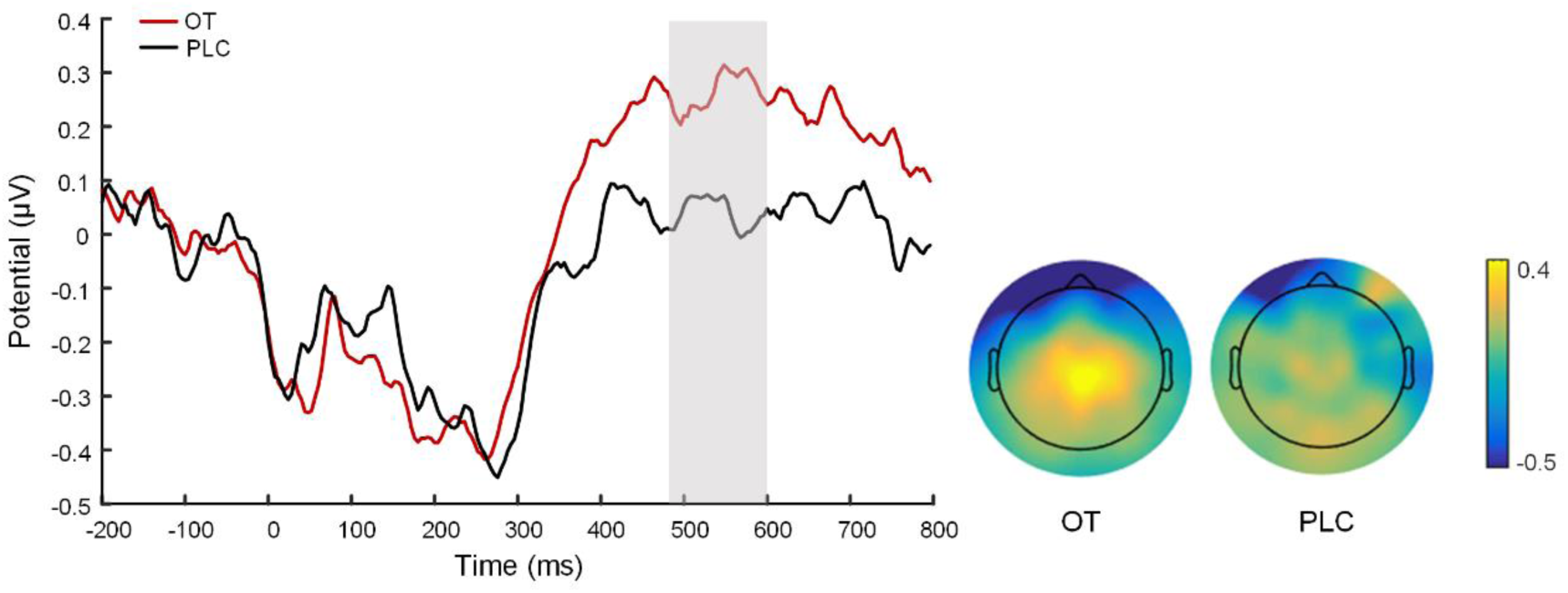
The HEP component and topographical maps at frontocentral and central electrodes following OT and PLC treatments in the heartbeat counting task.

#### Correlations

Pearson correlation analyses found a marginal positive correlation between HRV and IAc in the HCT following PLC (r = 0.333, *p* = 0.050) but not OT treatments (r = 0.209, *p* = 0.215; Fig. 4A). Fisher z-transformation test showed no significant correlation difference between the two groups (Fisher z-score = 0.544, *p* = 0.586). Furthermore, Spearman correlation analyses showed a significant positive correlation between IAc and HEP amplitudes across groups (r = 0.249, *p* = 0.035; Fig. 4B).

**Fig. 4.**
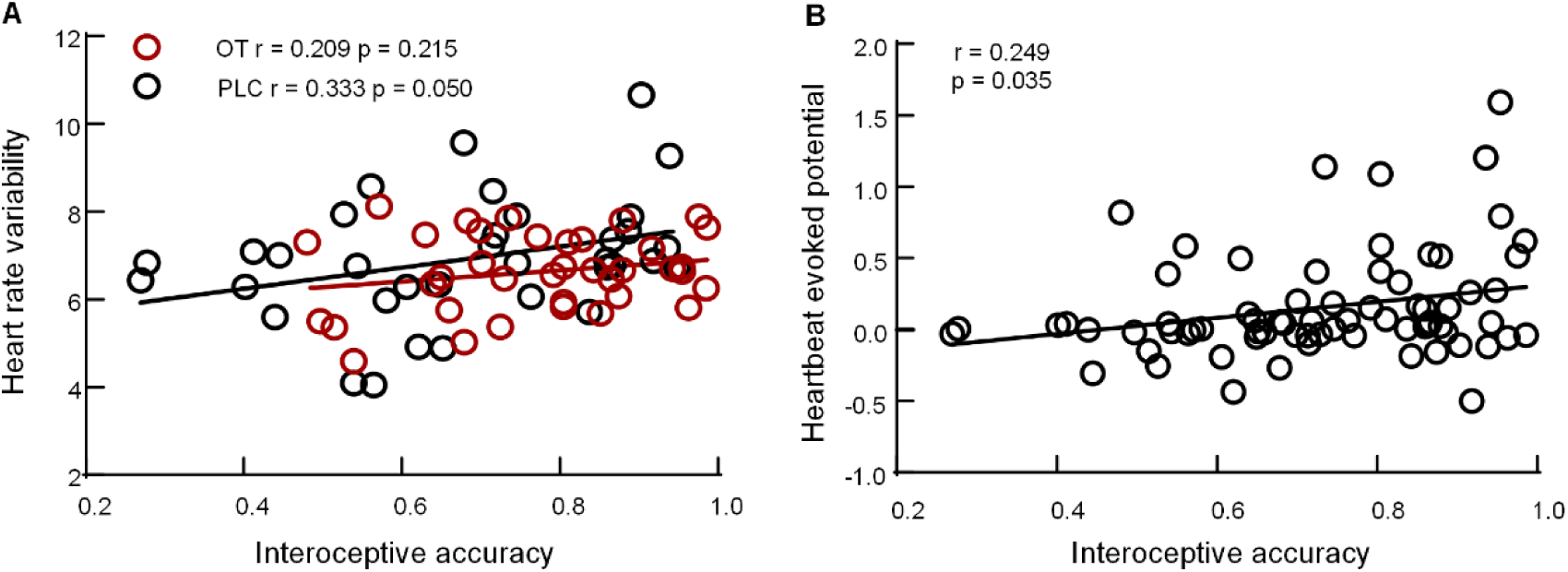
(A) A positive correlation between interoceptive accuracy and heart rate variability following PLC but not OT treatments. (B) A positive correlation between interoceptive accuracy and HEP amplitudes across treatment groups.

## Discussion

The present study recorded participants’ behavioral, ECG and EEG data related to cardiac interoceptive signals to investigate whether intranasal OT could modulate interoceptive processing. Our behavioral results revealed that OT significantly increased IAc compared to PLC in the HCT. This enhancement effect at the behavioral level was accompanied by lager HEP amplitudes following OT relative to PLC treatments on the neural level. HEP amplitudes were found to be positively correlated with IAc in the HCT across groups. Additionally, there was also a positive correlation between IAc and HRV in the PLC but not the OT group. However, we found no evidence for significant effects of OT on modulating the HR and HRV in both conditions or HEP amplitudes in the resting-state condition.

More specifically, participants in the OT group exhibited higher IAc in the HCT relative to the PLC group, suggesting that OT enhances the ability to perceive cardiac interoceptive signals in healthy participants. Consistent with these behavioral findings, at the neural level we found that OT significantly increased HEP amplitudes. Given that the HEP has been proposed as a neurophysiological marker of interoceptive processing (1,28), higher amplitudes of the HEP also suggest a faciliatory effect of OT on interoceptive processing. Although no studies have directly investigated effects of OT on task-based HEP, previous studies have demonstrated positive associations between HEP amplitudes recorded during the face affective-judgment and physical-judgment tasks and empathic concern scores (3). The HEP has also been found to show stronger responsivity to emotional video clips relative to neutral ones (30). Given previous studies demonstrating the effects of OT on improving empathy, emotional recognition, and self-referential bias (31,46–48) and previous findings of close associations between interoceptive processing and empathy, emotional recognition, and self-processing (49–52), it is not surprising that we observed a faciliatory effect of OT on interoceptive processing at both the behavioral and neural levels. Furthermore, we also found a positive correlation between IAc and HEP amplitudes in all samples in the HCT, which is in line with previous studies demonstrating that individuals with higher IAc show larger HEP than those with lower IAc (24,26). For IS, we did not find significant treatment effects in the HCT which is consistent with previous studies reporting the independence between IAc and IS in the HCT (26,33).

Note that the faciliatory effect of OT on interoceptive processing seems contradictory with previous findings particularly at the behavioral level. In healthy participants, Yao et al. (20) conducted a revised heartbeat detection task whereby participants were instructed to tap to indicate their heartbeats. Findings showed that while OT had no significant effects on IAc and related neural activity in the interoceptive network, when participants were simultaneously presented with social stimuli (neutral and emotional faces) OT decreased IAc, possibly by switching attention away from interceptive signals towards external salient social cues via enhancing the anterior insula responses and their control over the posterior insula. The divergence between findings in this previous study and the present one could be firstly due to different cardiac interoceptive paradigms used and secondly that Yao et al. (20) conducted the experiment during fMRI scanning where stronger noise may interfere with heartbeat perception compared to during EEG recording. Another study found that OT decreased IAc in the HCT in both heavy and light social drinkers but increased IAc only in heavy drinkers in the heartbeat discrimination task (integrating interoceptive and external signals) (19). Different participant populations and interoceptive paradigms thus may contribute to contradictory findings between the present study and Betka et al. (2018). These interpretations are also in accordance with previous findings that OT’s effects on human behavior can be influenced by the nature of contexts, individual differences, and experimental tasks (13,53).

To replicate previous findings, we also measured the HEP in the resting-state condition but found no modulatory effect of OT on resting-state HEP. This is consistent with a previous study which also reported an absence of effects of OT on the resting-state HEP response in both healthy controls and borderline personality disorder patients (29). We also measured HR and HRV responses in both resting-state and task-based conditions and showed no significant effects of OT on these two indices in the two conditions. These findings thus lend support for a null effect of OT on these two parasympathetic indices (54–56), although findings in this field are controversial given differences of within-vs. between-subject design and administered dosages of OT (57,58). The absence of effects of OT on HR also helps exclude the possible confounding effect of the HR on IAc in the HCT (26,59,60). Furthermore, a marginally positive correlation was found between IAc and HRV in the HCT following PLC treatment, which was in accordance with a previous study demonstrating that individuals with higher HRV showed higher IAc than those with lower HRV (61). This positive correlation was blunted by OT treatment possibly due to OT increasing IAc and thus inducing a right-skewed distribution of IAc as suggested by the scatter plot.

Several limitations should be acknowledged in the present study. First, only males were recruited and thus findings may not be generalizable to females. Second, only cardiac interoceptive signals were measured and we therefore cannot determine whether OT’s faciliatory effects can be extended to other modalities of interoceptive signals from such as respiration, intestines and stomach, which need to be explored in future studies. Third, the construct validity of precisely measuring IAc using the HCT is controversial since participants’ performance can be influenced by time estimation and prior knowledge of heartbeat (22,62,63). Although optimized instruction was used to minimize confounding effects from time estimation and prior knowledge of heartbeat (21,22), we still cannot completely exclude potential confounding effects. Future studies are needed to develop new optimal paradigms for measuring interoception.

In conclusion, the present study examined the impact of OT on the HEP as a neurophysiological marker of interoceptive processing and demonstrated an enhancement effect of OT on HEP amplitudes relative to PLC in the HCT. This neural effect is paralleled by a higher IAc following OT treatment on the behavioral level, with the IAc being positively correlated with HEP amplitudes. We also replicate previous findings of null effects of OT on resting-state HEP and cardiac responses. These findings together highlight a facilitatory effect of OT on cardiac interoceptive processing in the HCT. Our study not only provides new insights into interoceptive processing and its oxytocinergic modulation but also proof of concept evidence for the therapeutic potential of intranasal OT in disorders with dysfunctional interoception such as borderline personality disorder, major depressive disorder and obsessive-compulsive disorder (64–66).

## Data Availability

All data produced in the present study are available upon reasonable request to the authors

## Acknowledgements

This study was supported by the Natural Science Foundation of Sichuan Province (grant number: 2023NSFSC1187) and the Humanity and Social Science Foundation of Ministry of Education of China (grant number: 22XJC190003). We would like to thank all of the participants who gave their time and effort to this study.

